# Aerosolization of viable *Mycobacterium tuberculosis* bacilli by tuberculosis clinic attendees independent of sputum-GeneXpert status

**DOI:** 10.1101/2022.11.14.22282157

**Authors:** Benjamin Patterson, Ryan Dinkele, Sophia Gessner, Anastasia Koch, Zeenat Hoosen, Vanessa January, Bryan Leonard, Andrea McKerry, Ronnett Seldon, Andiswa Vazi, Sabine Hermans, Frank Cobelens, Digby F. Warner, Robin Wood

**Affiliations:** University of Amsterdam, Amsterdam Institute for Global Health and Development; Amsterdam, The Netherlands; SAMRC/NHLS/UCT Molecular Mycobacteriology Research Unit & DSI/NRF Centre of Excellence for Biomedical TB Research, Department of Pathology, Faculty of Health Sciences, University of Cape Town; Cape Town, South Africa; Institute of Infectious Disease and Molecular Medicine, Faculty of Health Sciences, University of Cape Town; Cape Town, South Africa; Desmond Tutu Health Foundation; Cape Town, South Africa; Wellcome Centre for Infectious Diseases Research in Africa, Faculty of Health Sciences, University of Cape Town; Cape Town, South Africa

## Abstract

The potential for bioaerosol release of *Mycobacterium tuberculosis* (*Mtb*) during different tuberculosis (TB) disease states is poorly understood. We quantified viable aerosolized *Mtb* from presumptive TB patients on diagnosis and, thereafter, through six months’ standard chemotherapy. At presentation, TB clinic attendees (n=102) were classified by laboratory, radiological, and clinical features into Group A: Sputum-GeneXpert-positive TB (n=52), Group B: Sputum-GeneXpert-negative TB (n=20), or Group C: TB not diagnosed (n=30). All were assessed for *Mtb* bioaerosol release at baseline, and subsequently at two weeks, two months, and six months. In Groups A and B, comprising notified TB cases, *Mtb* was isolated from 92% and 90% of participants at initial presentation; 87% and 74% at two weeks; 54% and 44% at two months; and 32% and 20% at six months, respectively. Surprisingly, similar numbers were detected in Group C: 93%, 70%, 48%, and 22% at the same timepoints. We also observed a temporal association between *Mtb* bioaerosol release and TB symptoms in all three groups, with 30% of participants remaining *Mtb* bioaerosol positive at six months irrespective of TB chemotherapy. Captured *Mtb* bacilli were predominantly acid-fast stain-negative and poorly culturable; however, following *in vitro* incubation, one sputum-GeneXpert-positive and two sputum-GeneXpert-negative aerosol samples yielded sufficient biomass for whole-genome sequencing, revealing two different *Mtb* lineages. The detection of viable aerosolized *Mtb* in most clinic attendees at presentation, independent of TB diagnosis, suggests that unidentified *Mtb* transmitters could account for a significant attributable proportion of community exposure. However, longitudinal studies are required to investigate this possibility.

**One Sentence Summary:** *M. tuberculosis* bacilli are detected in bioaerosols of presumptive tuberculosis (TB) patients irrespective of final TB diagnosis and clear over time.

## INTRODUCTION

Tuberculosis (TB) is uniquely characterized by obligate airborne transmission (*1*) with each new infection initiated following deposition of small diameter (<5μm) *Mycobacterium tuberculosis* (*Mtb*)-containing bioaerosols in the distal lung of a susceptible human host (*2*). *Mtb* bioaerosols originate from infected individuals, and proximity to potential new hosts is necessary for rebreathing of viable organisms to occur. The onward propagation of TB therefore depends on the processes of natural production, expulsion, and survival of airborne *Mtb* in small aerosols. We recently reported considerable variability in cough-independent release of particulate matter – including *Mtb* organisms – among confirmed TB patients (*3*). However, the influence of TB disease stage on individual capacity for *Mtb* bioaerosol release is not known. Similarly, the speed and impact of standard TB chemotherapy on *Mtb* aerosolization has not been directly quantified.

The lung has been described as an aerosol generator (*4*): every exhaled breath contains a small volume of peripheral lung fluid released by a fluid film burst mechanism in terminal bronchioles (*5*). Bioaerosol sampling captures this peripheral lung contribution and can be thought of as a non-invasive broncho-alveolar lavage. In contrast, sputum – which is produced in disease states – derives from mucin hyperproduction by goblet cells located in the upper conducting airways and confined to the trachea, bronchi, and larger bronchioles (*6*). Bioaerosols might, therefore, represent a more suitable sample to identify *Mtb* in the lung periphery, and to investigate individuals with no or minimal sputum production, including subclinical TB disease (*7*). Notably, this sample has value both in demonstrating the presence of pathogen (*Mtb*) and, potentially, indicating a quantitative transmission risk.

Given the low airborne bacillary load, bioaerosol investigations depend on efficient capture and high sensitivity detection of viable *Mtb* (*8*). Previous studies have explored respiratory maneuvers (*9*), collection devices (8), and the proximity of the participant to the collecting apparatus (*10*) to optimize capture of aerosolized *Mtb*. Groups employing cough aerosol (*11*) or facemask (*12*) sampling systems have achieved sensitivities of 30% and 91%, respectively, in sputum-positive patients. Combining advanced aerosol sampling and fluorescence detection systems, we have previously reported a 95% yield of viable *Mtb* in sputum-GeneXpert-positive individuals (*13*).

Utilizing the same technology, we aimed in the current study to investigate the capacity for viable *Mtb* release by TB clinic attendees categorized into three mutually exclusive groups based on South African National TB Program treatment and diagnostic protocols (*14*): laboratory-confirmed (sputum-GeneXpert-positive) TB (Group A), sputum-GeneXpert-negative TB (Group B), and those not diagnosed with TB (Group C). By sampling all participants at defined intervals (two weeks, two months, and six months) post initial presentation, we further aimed to determine the impact of standard TB chemotherapy on *Mtb* bioaerosol production. The results presented here reveal the unexpected production of *Mtb* bioaerosols by non-TB patients, a time-dependent (not drug-dependent) decline in bioaerosol positivity, and the sustained detection of *Mtb* bioaerosols in a proportion of confirmed TB patients despite completion of the standard six-month combination regimen.

## RESULTS

### Demographic and symptomatic characteristics of the study cohort

We recruited consecutive presumptive pulmonary TB patients over the age of 13 who self-presented to two community clinics serving two high-density, peri-urban residential areas south-west of Cape Town, South Africa. The final study population consisted of 102 TB clinic attendees (Table 1) who underwent repeated bioaerosol sampling between 15 May 2020 and 27 May 2022.

**Table 1.** Characteristics of patients stratified by each of the three diagnostic groups. Normally distributed continuous variables are reported as means with standard deviations (SD) in brackets. Medians are reported with interquartile range (IQR) values. Categorical variables are reported as counts with percentages in brackets.

| **Group** | **A**<br>**Sputum-GeneXpert-**<br>**Positive** | **B**<br>**Sputum-GeneXpert-**<br>**Negative** | **C**<br>**TB Not**<br>**Diagnosed** |
| --- | --- | --- | --- |
| **n** | 52 | 20 | 30 |
| **Age (mean (SD))** | 33.6 (9.8) | 45.0 (10.9) | 39.6 (11.7) |
| **Sex, Male (%)** | 32 (61.5) | 12 (60.0) | 19 (63.3) |
| **BMI (mean (SD))** | 19.9 (3.9) | 20.6 (2.9) | 23.9 (6.3) |
| **HIV status (%)** |  |  |  |
| **Negative** | 33 (63.5) | 5 (25.0) | 18 (60.0) |
| **Positive** | 19 (36.5) | 15 (75.0) | 11 (36.7) |
| **Unknown** | 0 (0.0) | 0 (0.0) | 1 (3.3) |
| **On ARVs (%)** |  |  |  |
| **Defaulted before the study** | 9 (50.0) | 5 (35.7) | 0 (0.0) |
| **No** | 4 (22.2) | 3 (21.4) | 3 (27.3) |
| **Yes** | 5 (27.8) | 6 (42.9) | 8 (72.7) |
| **CD4 Count (median [IQR])** | 162 [95, 245] | 144 [58, 214] | 148 [86, 242] |
| **On INH prophylaxis (%)** |  |  |  |
| **Defaulted before the study** | 1 (2.2) | 1 (5.3) | 0 (0.0) |
| **No** | 43 (95.6) | 18 (94.7) | 21 (91.3) |
| **Yes** | 1 (2.2) | 0 (0.0) | 2 (8.7) |
| **Previous TB (%)** | 17 (32.7) | 13 (65.0) | 15 (50.0) |
| **No. of Previous TB Episodes (%)** |  |  |  |
| **1** | 11 (61.1) | 8 (66.7) | 13 (86.7) |
| **2** | 3 (16.7) | 2 (16.7) | 2 (13.3) |
| **3** | 4 (22.2) | 2 (16.7) | 0 (0.0) |
| **Pre-existing Lung Disease (%)** | 3 (5.9) | 0 (0.0) | 4 (13.3) |
| **Smoking (%)** | 25 (51.0) | 10 (52.6) | 10 (33.3) |

BMI, body mass index; ARV, anti-retroviral; INH, isoniazid.

Of the participants sampled, 52 were sputum-GeneXpert-positive (Group A); 20 were diagnosed as sputum-GeneXpert-negative TB (Group B), 19 based on chest radiography and one on clinical suspicion alone; and 30 were sputum-GeneXpert-negative and not diagnosed with TB during six months of follow-up visits (Group C). One participant initially categorized as Group C was reclassified to Group B following radiographic TB diagnosis after the second visit. Notified TB patients (Groups A and B) received standard short-course chemotherapy for six months comprising 2 months’ rifampicin, isoniazid, pyrazinamide, and ethambutol combination therapy, followed by 4 months’ rifampicin plus isoniazid. Aerosol sampling at the initial visit was performed immediately before initiation of TB treatment in those diagnosed with TB. Eight participants in Group C received short-course amoxicillin at initial presentation; no quinolones were prescribed. Through the study, 16 participants were lost to follow-up, four relocated, four refused further sampling, three died, and one presented with multidrug-resistant TB and was excluded. In total, there were 352 sampling visits with 72 participants completing all four visits: baseline, two weeks, two months, and six months post initial presentation.

There were some differences in the baseline characteristics of the three groups (Table 1): Group A recorded the lowest median age and prevalence of previous TB, Groups A and B had lower body mass indices (BMIs) than Group C, and individuals in Group B were older and had a higher prevalence of HIV infection. Notably, 45 participants (44.1%) had a history of previous TB.

At presentation (baseline sample), nearly all participants reported at least one symptom (Fig. 1A). The median number of symptoms was highest in Group A (Fig. 1B), as expected; however, all three groups reported weight loss, persistent cough, loss of appetite, and night sweats as the most prevalent symptoms (Table 2).

**Fig. 1.**
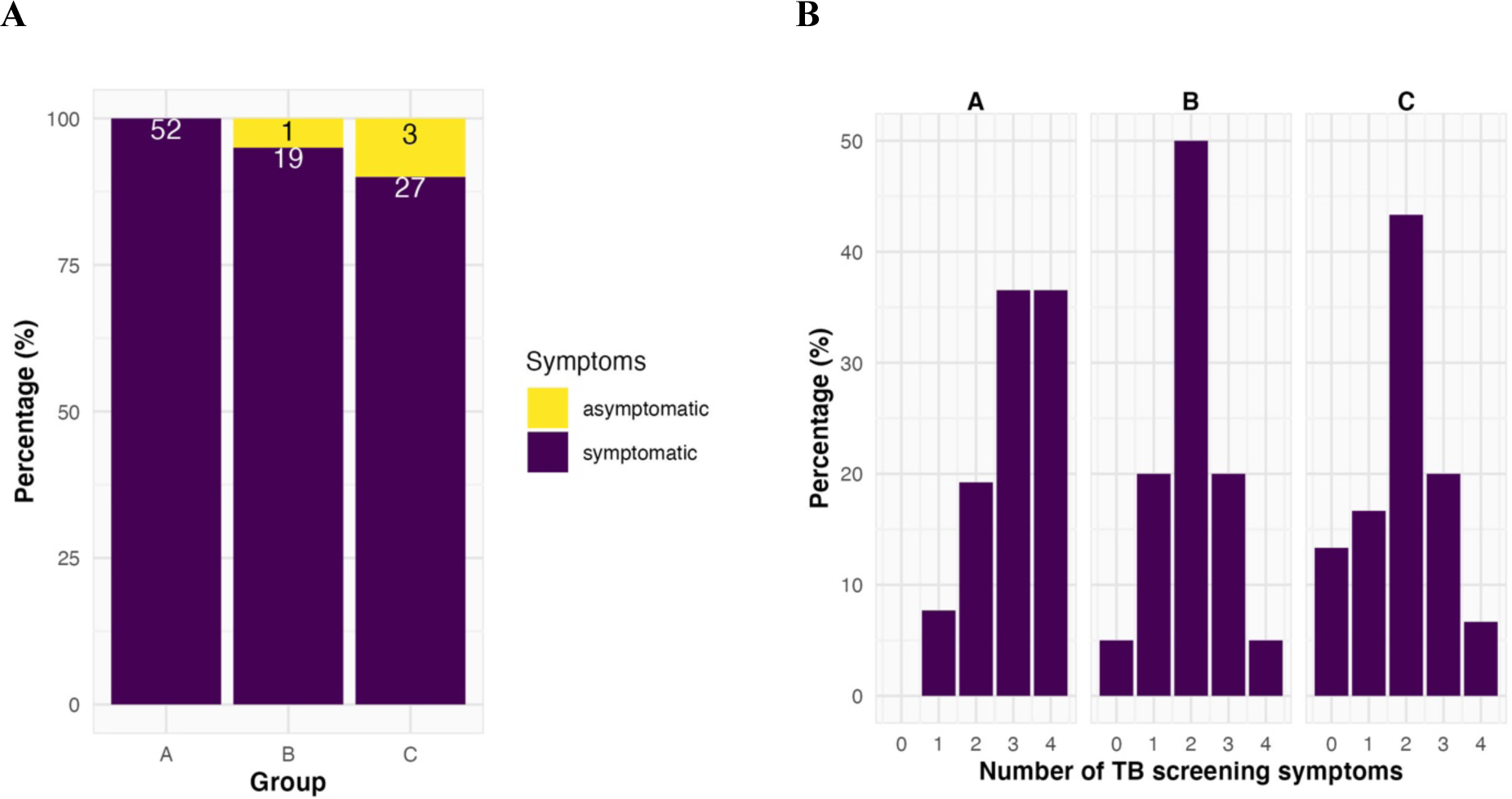
Presentation of symptoms at baseline. (**A**) Percentage of participants with one or more self-reported symptom. (**B**) Distribution of the number of symptoms per individual by diagnostic group; the median symptom number for Group A, B, and C was 3, 2, and 2, respectively.

**Table 2:** The prevalence of each of the symptoms identified by patient questionnaire at the initial visit stratified by diagnostic group.

| Group | A<br>Sputum-GeneXpert-Positive | B<br>Sputum-GeneXpert-Negative | C<br>TB Not<br>Diagnosed |
| --- | --- | --- | --- |
| n | 52 | 20 | 30 |
| **Persistent Cough (%)** | 45 (86.5) | 12 (60.0) | 19 (63.3) |
| **Weight Loss (%)** | 46 (88.5) | 16 (80.0) | 20 (66.7) |
| **Night Sweats (%)** | 38 (73.1) | 8 (40.0) | 12 (40.0) |
| **Loss of Appetite (%)** | 28 (53.8) | 4 (20.0) | 6 (20.0) |
| **Fever (%)** | 7 (13.5) | 1 (5.0) | 8 (26.7) |
| **Haemoptysis (%)** | 5 (9.6) | 0 (0.0) | 2 (6.7) |
| **Myalgias (%)** | 8 (15.4) | 1 (5.0) | 4 (13.3) |
| **Anosmia (%)** | 1 (1.9) | 1 (5.0) | 0 (0.0) |

### Microbiological detection of Mtb in bioaerosol samples

Each participant produced bioaerosols from three distinct respiratory maneuvers – force vital capacity (FVC), tidal breathing, and voluntary cough. DMN-trehalose-positive *Mtb* bacilli with characteristic morphological appearance were detected for all three diagnostic groups. No significant differences were observed in the numbers of *Mtb* bacilli detected nor the prevalence of positive samples between the three respiratory maneuvers (Figs. S1A & B). Some individuals displayed a greater propensity for bacillary aerosolization in a single respiratory maneuver, however this was not consistent for all participants tested (Fig. S1C). Consequently, counts from respiratory maneuvers were pooled in subsequent analyses to compare participants in the three diagnostic categories, Groups A-C.

Surprisingly, despite the differences reported in symptom number and severity, we observed no significant differences in median *Mtb* counts across Groups A-C at baseline (Fig. 2A). Moreover, the prevalence of *Mtb*-containing bioaerosols was similar for all three groups (Fig. 2B), with *Mtb* detected in 92%, 90%, and 93% of bioaerosol samples in Groups A, B, and C, respectively. There were no clinical or demographic variables predictive of a positive *Mtb* bioaerosol (Table 3).

**Fig. 2.**
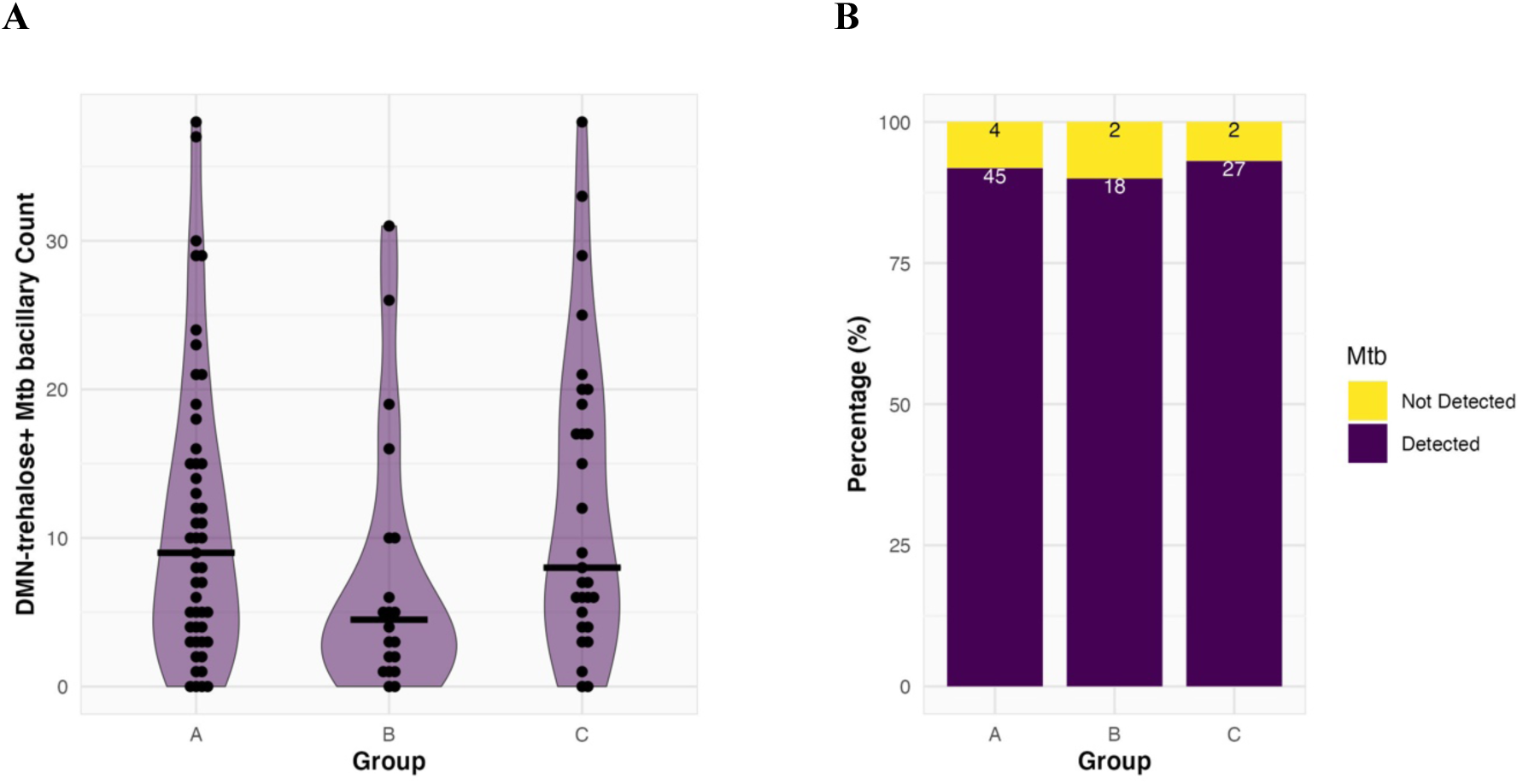
The detection of *Mtb* bacilli in the bioaerosol of all three diagnostic groups at baseline. (**A**) Counts of putative *Mtb* with medians 9, 4.5 and 8 (represented by the thick black bars) for Group A, B and C respectively. No difference was found between the groups with Wilcoxon Rank Sum testing after Bonferroni correction for multiple comparisons. (**B**) Percentage *Mtb* bioaerosol positive samples per diagnostic group. Owing to contamination of the slides, 4 samples were uninterpretable; these are not included in the denominator of the prevalence proportions

**Table 3.** Odds ratios for baseline demographic and clinical variables associated with baseline DMN-trehalose *Mtb* bacillary positivity.

|  | **Unadjusted**<br>**Odds Ratio** | **CI low**<br>**(2.5)** | **CI high**<br>**(97.5)** | **P-value** |
| --- | --- | --- | --- | --- |
| **Age** |  |  |  |  |
| <30 | *Reference* |  |  |  |
| ≥30 | 1.84 | 0.26 | 10.37 | 0.443 |
| **Sex** |  |  |  |  |
| Male | *Reference* |  |  |  |
| Female | 1.81 | 0.3 | 19.33 | 0.514 |
| **Body Mass Index** |  |  |  |  |
| <20 | *Reference* |  |  |  |
| ≥20 | 0.15 | 0 | 1.24 | 0.053 |
| **HIV status** |  |  |  |  |
| Negative | *Reference* |  |  |  |
| Positive | 0.82 | 0.14 | 4.69 | 0.792 |
| **GeneXpert** |  |  |  |  |
| Negative | *Reference* |  |  |  |
| Positive | 1 | 0.17 | 5.72 | >0.99 |
| **Isoniazid Preventive Therapy\*** |  |  |  |  |
| No | *Reference* |  |  |  |
| Yes | 0.27 | 0.03 | 14.3 | 0.772 |
| **Previous TB** |  |  |  |  |
| No | *Reference* |  |  |  |
| Yes | 1.45 | 0.26 | 9.91 | 0.647 |
| **Smoking History** |  |  |  |  |
| No | *Reference* |  |  |  |
| Yes | 1.55 | 0.28 | 10.59 | 0.589 |
| **Underlying Lung Disease\*** |  |  |  |  |
| No | *Reference* |  |  |  |
| Yes | 0.67 | 0.08 | 29.13 | 0.54 |

\*Haldane-Anscombe correction used for small samples.

### Secular trends in the detection of Mtb in bioaerosol samples

In compliance with SA National TB Control Program policy, sputum-GeneXpert-positive (Group A) and clinically diagnosed sputum-GeneXpert-negative (Group B) TB patients were immediately started on six months’ standard anti-TB therapy. Owing to the unexpectedly high baseline identification of aerosolized viable *Mtb* across all groups (A-C), we decided to monitor all participants for *Mtb* bioaerosol release and TB symptoms for six months at defined intervals, regardless of clinical TB diagnosis. Therefore, serial bioaerosol sampling was conducted at approximately two weeks (a two week timepoint was targeted but, in practice, the median was 19 days, IQR 10 days), two months (median 61 days, IQR 19 days), and six months (median 176 days, IQR 27 days) after initial enrolment (Fig. S2).

*Mtb* bioaerosol numbers and the percentage of *Mtb*-positive bioaerosol samples decreased equivalently in all three groups over the six months, irrespective of TB therapy (Fig. 3A). In Groups A and B – comprising notified TB cases – MTB was isolated in bioaerosol samples from 87% and 74% of participants at two weeks, 54% and 44% at two months, and 32% and 20% at six months, respectively, with similar numbers detected for Group C (70%, 48% and 22%, respectively, at the same timepoints). Notably, the decline in bioaerosol positivity corresponded with a reduction in symptoms for all three groups (Fig. 3B), and there were no notable differences in the numeric trends between any of the three groups (Fig. S3). Furthermore, no baseline predictors of *Mtb* bacillary clearance were identified at any of the timepoints (Fig. S4). The median time to clearance of *Mtb* bacilli from bioaerosol samples was 83 days (95% CI, 63 to 167 days) (Fig. 4A); stratification by treatment (Fig. 4B), previous history of TB (Fig. 4C), or HIV status (Fig. 4D), did not significantly alter the time to clearance (log-rank p-values 0.751, 0.404, and 0.798 respectively). The median time to TB symptom resolution was 168 days (95% CI, 166 to 176 days) (Fig. 5A). A previous history of TB (Fig. 5C) was associated with persistence of symptoms towards the end of the observation period for a minority of participants (log rank p-value, 0.026). There was no impact of treatment (Fig. 5B) nor HIV status (Fig. 5D) on rate of symptom resolution (log-rank p-values, 0.210 and 0.497, respectively).

**Fig. 3:**
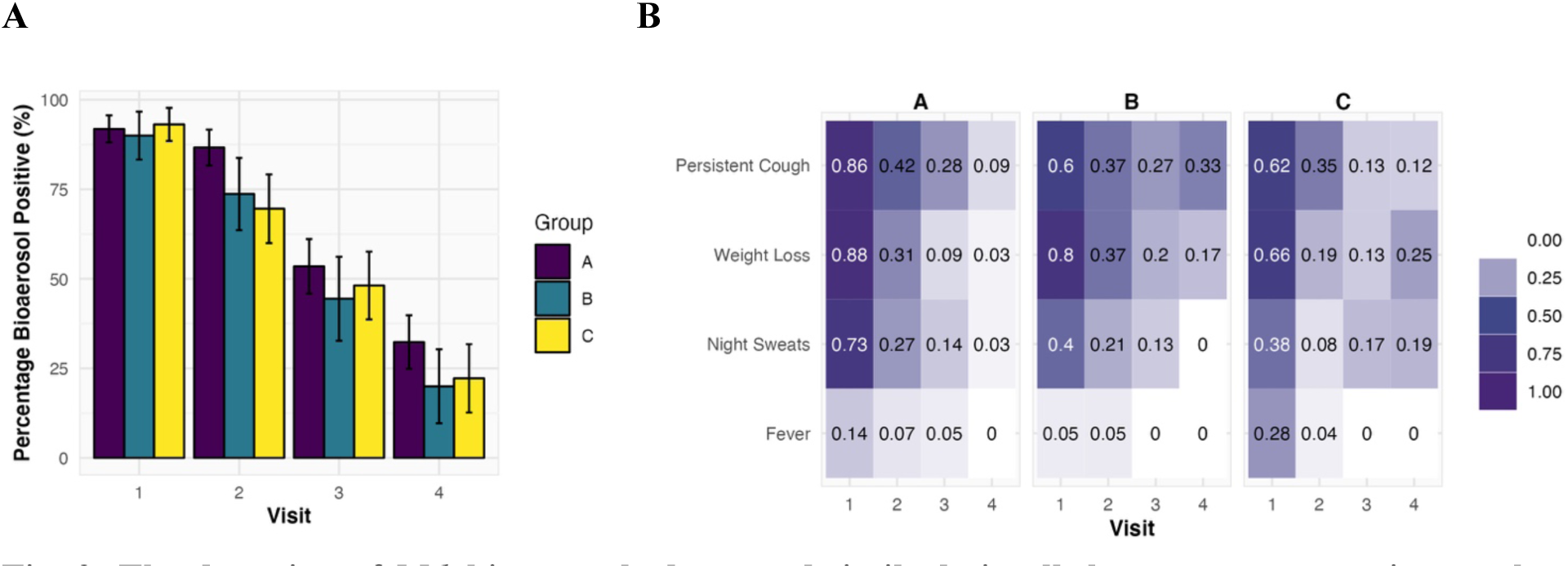
The detection of *Mtb* bioaerosols decreased similarly in all three groups over six months. **(A)** The percentage of bioaerosol samples that were positive for *Mtb*. The denominator is all visits yielding an interpretable sample. 13 samples were uninterpretable; these are not included in the denominator of the prevalence proportions. The error bars represent the standard error of the mean. Fisher’s exact tests performed on each visit found no significant differences. (**B**) A heat map indicating the prevalence of TB screening symptoms across visits. The numbers indicate the symptom proportion of patients in each group-visit combination with symptoms.

**Fig. 4.**
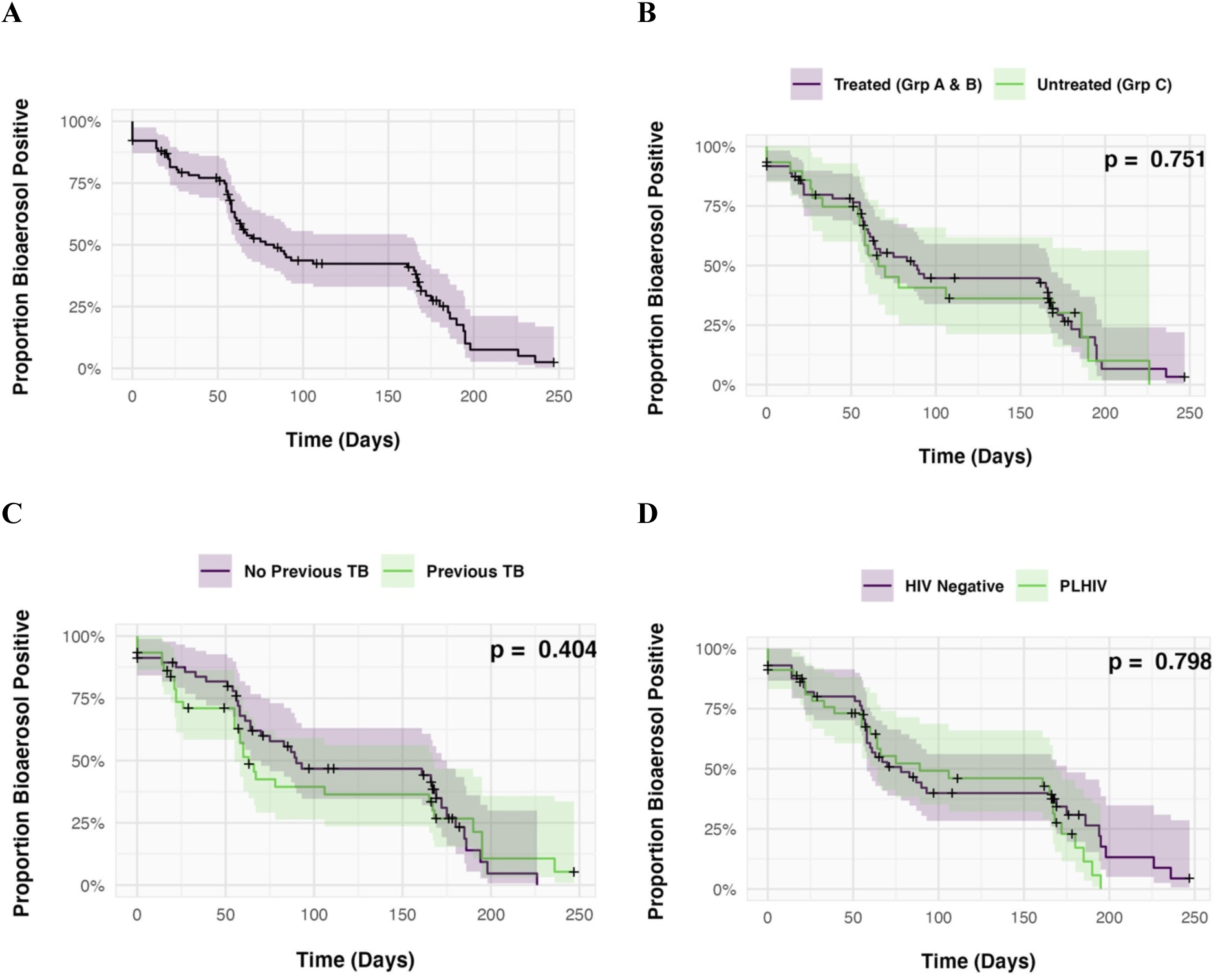
Kaplan-Meier plots showing time to *Mtb* bacillary clearance from collected bioaerosol. The first visit at which *Mtb* is not detected in the sample is considered the time to clearance. Individuals with detectable *Mtb* at the final visit are censored at the time of this visit. (**A**) All participants. (**B**) TB patients on treatment (Groups A & B) are separated from those individuals not diagnosed with TB and therefore not treated (Group C). (**C**) Comparison of time-to-clearance between those with a previous history of TB and those without. (**D**) Participants separated according to HIV status. The shaded areas represent 95% confidence intervals. Log rank test p-values are given in the top right corner for panels B, C and D.

**Fig. 5.**
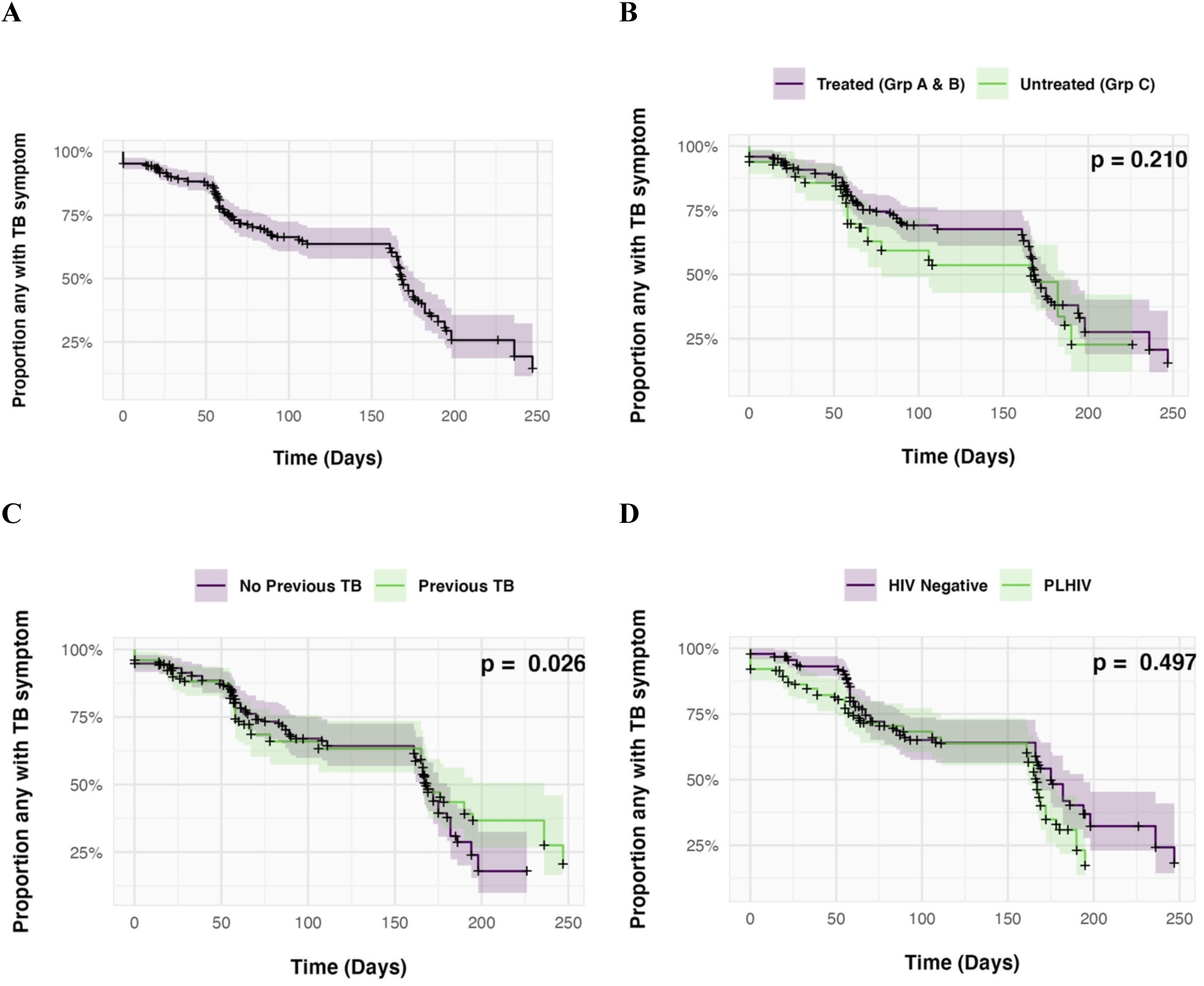
Kaplan-Meier plots showing the proportion of individuals with any TB symptom (persistent cough, fever, weight loss, night sweats) against days since first sampling. The first visit without reported symptoms is considered the time to clearance. Participants with symptoms at the final visit are censored at time of this visit. (A) All participants. (B) Participants separated into TB patients on treatment (Group A, B) and those not diagnosed with TB and therefore not treated (Group C). (C) Comparison of participants with a previous history of TB and without. (D) Comparison of participants according to HIV status. The shaded areas represent 95% confidence intervals. Log rank test p-values are given in the top right corner for panels B, C and D.

To evaluate the possibility of a change in DMN-trehalose assay sensitivity over time, DMN-trehalose *Mtb* bacilli counts were assessed for correlation with calendar date for each group and visit combination. The proportion of the variation explained by time was a median of 3% (R-squared range: <0.01 to 0.15).

### Microscopic characterization of Mtb bacilli from bioaerosol samples

Mycobacteria, including *Mtb*, demonstrate morphological and metabolic plasticity in response to environmental and antibiotic stresses (*15–17*). We reasoned that, if *Mtb* were exposed to antibiotics before aerosolization, a discernible change in bacterial morphology and/or metabolic state might be detectable between treatment groups. An important consideration before performing this analysis, however, was the previous observation that *Mtb* morphology, specifically cell length, can differ according to anatomical origin (*18*). To allow for data pooling, we first needed to confirm that the three maneuvers employed in this study produced *Mtb* from the same respiratory compartment. To this end, we compared the length distributions of *Mtb* detected from each respiratory maneuver at baseline (Fig. S5). No significant differences were evident, suggesting that all bacilli were aerosolized from the peripheral lung by a conserved mechanism (*3,9*).

As observed previously (*19*), we identified multiple DMN-trehalose staining profiles in *Mtb* bacilli captured from participant bioaerosols (Figs. S6A & S6B), with most baseline samples characterized by two or more distinct staining patterns (Fig. S6C). For further microscopic analyses, the six month sample was excluded as too few bacilli were detected to make reliable conclusions.

Polarity index, a metric summarizing the staining profile of synthetic trehalose probes along the medial axis, has previously been shown to differ significantly upon exposure to antibiotics (*20*). Comparing the average polarity index of bacilli from treated and untreated samples did not reveal significant differences at baseline or two weeks. However, the polarity index of *Mtb* from treated individuals was slightly higher at two months (Fig. 6A) and corresponded with a minor increase in cell length observed in the same samples (Fig. 6B). Despite these very slight changes in cell length and DMN-trehalose staining, no significant differences in clustering were observed between participants (Fig. S6), suggesting that the treatment effect on bacterial phenotype was minor.

**Fig. 6.**
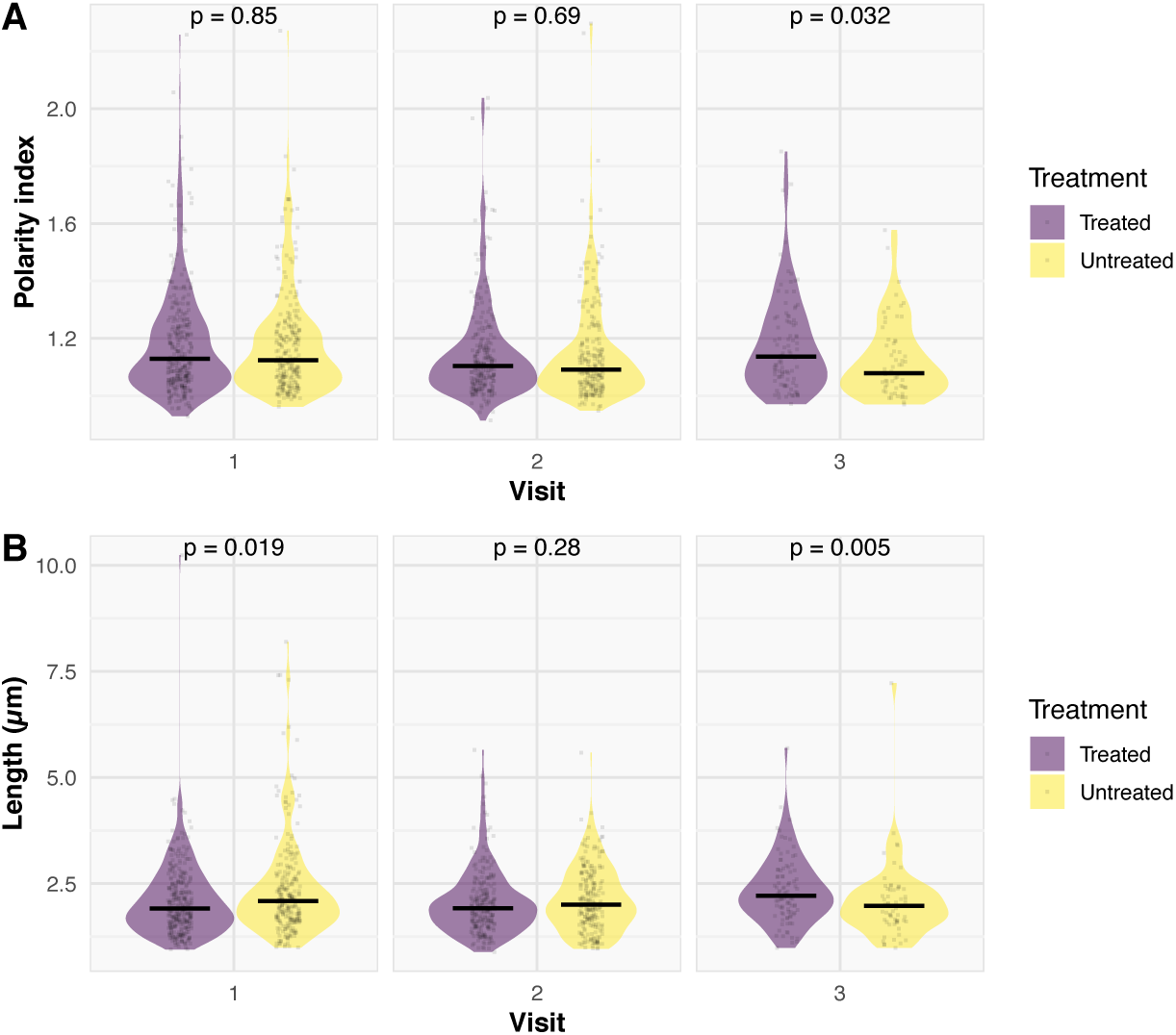
Examining the change in DMN-trehalose phenotype and *Mtb* cell length through time. A comparison of (**A**) polarity index and (**B**) *Mtb* cell length between treatment groups at each visit. Polarity index is defined as the fluorescence intensity in arbitrary fluorescence units (AFU) of the brighter pole divided by the fluorescence intensity at the mid-cell. A Wilcoxon Rank Sum test was performed.

### Genotypic and genomic confirmation of Mtb in bioaerosol samples

The apparent lack of a treatment effect on *Mtb* bioaerosol clearance was a surprising observation. To increase our confidence in the *Mtb* assignments based on DMN-trehalose fluorescence, a second bioaerosol sample was collected in parallel from a subset of participants. The presence of *Mtb* bacilli in these samples was assessed using either conventional Auramine O (n=44) and Ziehl-Neelsen (n=50) staining, or by means of a droplet digital (dd)PCR-based molecular assay targeting the *Mtb*-specific RD9 locus (n=40), as reported previously (*7*). Sequentially selected samples included both baseline and subsequent visits. All (40/40) of the samples investigated by ddPCR were RD9 positive, confirming the presence of *Mtb* genomes. In contrast, Auramine O staining was positive in 82% (36/44) of samples, while Ziehl-Neelsen acid-fast staining was negative for all samples assayed.

Attempts to culture *Mtb* organisms from bioaerosol samples proved mostly unsuccessful despite extending the incubation period to 50 days in liquid medium. Five samples which exhibited some growth were selected for whole-genome sequencing to ascertain the feasibility of detecting “transmitted” *Mtb* lineages using this approach (Table 4): two from sputum-GeneXpert-positive (Group A) participants, one baseline and one at six months, and three from sputum-GeneXpert-negative (Group C) participants, two at baseline and one at two months. Of these, only three samples provided data of sufficient quality to detect lineage-defining SNPs (*21*). On analysis, we determined that one sputum-GeneXpert-positive sample (Group A, baseline) and one sputum-GeneXpert-negative sample (Group C, baseline) belonged to lineage 4.9, with the third sample (Group C, baseline) belonging to lineage 4.3 (Table 4).

**Table 4.** Bioaerosol samples (n=5) selected for *Mtb* whole-genome sequencing. Parallel samples shown by group and visit number. All samples were from separate individuals. *Mtb* DNA copy number determined by ddPCR for the *Mtb*-specific RD9 target is shown after 50-day incubation. *Mtb* lineage was determined for three samples.

| Sample | Group | Visit | *Mtb* DNA copies | *Mtb* Genome Coverage (fold) | Lineage |
| --- | --- | --- | --- | --- | --- |
| 1 | C | 1 | 38645 | 38.88 | 4.9 |
| 2 | A | 1 | 24793 | 30.30 | 4.9 |
| 3 | C | 1 | 1028 | 0.39 | 4.3 |
| 4 | A | 3 | 308 | 0.02 | ND |
| 5 | C | 4 | 89 | 0.06 | ND |

ND, Not determined.

## DISCUSSION

Current approaches to TB control are predicated on tenets which are increasingly being questioned as improved technologies enable insights into mycobacterial pathogenicity and the TB disease cycle that were previously inaccessible to investigation (*22*). Among these are the reliance on sputum *Mtb* positivity as a marker of patient infectiousness (*23*), and the dependence on coughing as sole vehicle for aerosol *Mtb* release (*3, 9, 13*). Historically, these assumptions have constrained most TB transmission research to the identification and recruitment of sputum-GeneXpert-positive or sputum-smear-positive TB patients, even in those utilizing innovative approaches to identify the “missing” TB cases, such as household contact investigations, and cough (*11*) or face-mask (*12*) sampling studies.

We previously reported the combined use of the RASC, DMN-trehalose labelling, and fluorescence microscopy to identify viable aerosolized *Mtb* in most sputum-positive TB patients (*10, 13, 24*). The DMN-trehalose probe is incorporated into the mycobacterial cell wall by the antigen-85 mycolyl transferase complex (*25*); detection of a fluorescent DMN-trehalose signal in a whole mycobacterial cell therefore indicates a metabolically active organism (*26*). Previous studies of bacillary size, morphology, and staining characteristics have shown good agreement between patient samples and cultures of the laboratory strain, *Mtb* H37Rv (*19*). DMN-trehalose staining has also demonstrated characteristic bacillary morphological and staining phenotypes that reliably distinguish *Mtb* from non-mycobacteria (*19*). Given that most infections cannot be linked to an index case (*27*), we hypothesized that *Mtb* might be aerosolized by some sputum-GeneXpert-negative individuals who would, by definition, be excluded from conventional TB transmission studies. Therefore, the primary aim of the current work was to determine the capacity for bioaerosol *Mtb* release by sputum-GeneXpert-negative TB patients (Group B) diagnosed on clinical presentation. By sampling all participants at defined intervals thereafter, we also aimed to ascertain the impact of standard TB chemotherapy on *Mtb* bioaerosol production.

We report two striking observations. First, we identified aerosolized *Mtb* in a large majority of both confirmed TB patients (sputum-GeneXpert-positive [Group A] and sputum-GeneXpert-negative [Group B]) and individuals excluded from a TB diagnosis (Group C) at initial clinic visit. In compliance with South African National TB Control Program policy, sputum-positive and clinically diagnosed sputum-negative TB patients immediately commenced 6 months’ standard anti-TB therapy. And, owing to the high baseline identification of aerosolized viable *Mtb*, we monitored all participants – regardless of clinical TB diagnosis – for aerosolized *Mtb* and TB symptoms at defined intervals for 6-months. This enabled the second major observation, namely that *Mtb* bioaerosol numbers and proportion positivity declined at similar rates irrespective of TB therapy. Notably, around 20% of all participants were aerosol positive at the 6-month timepoint, albeit with very low *Mtb* bacillary numbers.

The unexpected detection of *Mtb* in bioaerosols from individuals without proven TB challenged us to confirm the *Mtb* assignation utilizing alternative (DMN-trehalose-independent) approaches, and to better define the phenotypes of bioaerosol *Mtb*. To further address specificity, a series of secondary aerosol samples collected at the same visit was analyzed by ddPCR targeting the RD9 locus, an *Mtb*-specific genetic marker; all samples were RD9 positive, confirming the presence of *Mtb* genomic DNA. A large proportion of aerosol samples were also positive under Auramine O staining, which is routinely applied for sputum-based *Mtb* identification, but they were acid-fast stain-negative using the Ziehl-Neelsen protocol. The superior sensitivity and specificity of Auramine O over Ziehl-Neelsen staining has been reported previously (*28*) and has been ascribed to the retention of Auramine O by the mycolic acid component of the mycobacterial cell wall (*29*) – a conclusion which supports the correlation between Auramine O and DMN-trehalose positivity. Notably, loss of acid-fastness occurs naturally in human TB disease and animal infection models (*30*), and has been attributed to changes in cell wall composition and/or architecture as a function of metabolic alterations during host colonization (*29*). Whether aerosol bacilli are specifically adapted to external passage and survival is uncertain; however, the complete absence of acid-fast organisms in these samples raises important questions about the physiological state(s) of transmitted *Mtb* organisms.

Five samples that had been cultured for 50-days in 7H9 OADC media were selected for analysis via WGS to determine whether WGS could further validate the detection of genomes in bioaerosol samples, and more, if WGS data could provide information about the *Mtb* lineages observed. Three samples provided WGS data for which lineage associated SNPs could be reliably detected (Table 4). Two samples were classified as belonging to lineage 4.9 and one to lineage 4.3. Importantly, the samples that provided useful WGS data yielded >1000 DNA copies as estimated by RD9 ddPCR. These data indicate that while WGS is feasible on bioaerosols, sufficient DNA is required to generate data of high enough quality to identify specific *Mtb* lineages. Future work to facilitate WGS of a broader number of samples could include optimization of DNA extraction techniques, culture and possibly *Mtb* DNA enrichment prior to WGS (31–33).

A phenotype of poorly replicating, non-acid-fast *Mtb* able to cause TB in animal models with reversion to acid-fast staining was first identified from TB patients in 1883 (*34*). Subsequently isoniazid inhibition of mycolic acid biosynthesis demonstrated that cell wall organizational changes can be responsible for loss of acid-fast staining (*35*) and actively replicating acid-fast *Mtb* have been shown to convert to poorly replicating *Mtb* with an associated loss of acid-fast staining (*29*). The aerosol *Mtb* samples were poorly culturable, with only four of 64 (6.25%) yielding >1000 DNA copies after 50 days liquid culture. Poor culturability is a well-known characteristic of *Mtb* derived from clinical specimens (*36*) with at least one prior study reporting that ∼80% of sputum smear-positive samples required supplementation with resuscitation-promoting factor to stimulate growth (*37*). Very recent work involving single-cell analyses of *Mtb* organisms in microfluidic devices has also utilized spent culture medium to ensure reproducible growth (*38*), suggesting that future bioaerosol studies should incorporate this approach.

The finding of *Mtb* bacilli by non-invasive sampling of pulmonary lining fluid (PLF) from the lung periphery is compatible with the high diagnostic yield of bronchoalveolar lavage in sputum-negative TB (*39*) and consistent with previous evidence of transmission from this sub-group (*40–41*). The identification of viable *Mtb* in a subset of TB patients at EOT echoes previous findings of an FDG-PET/CT imaging study that tracked 99 HIV-negative patients through TB treatment and reported a range of outcomes with a proportion showing new FDG-avid lesions and ongoing inflammation (*42*). In that work, *Mtb* mRNA transcripts were found in 37% of sputum and 100% of bronchoalveolar lavage (BAL) samples from EOT patients, implying presence of live organisms. A further study of non-sterilizing cure found differentially culturable *Mtb* organisms in induced sputum and BAL after EOT, the presence of which correlated with FDG-PET/CT findings and subsequent clinical outcomes (*43*).

An important complementary finding from our study was that the rate of clearance of aerosolized *Mtb* appeared to be unaffected by treatment (Fig. 4B). This conclusion was supported by the observation that treatment did not significantly impact the morphology of bacteria from the bioaerosol (Fig. 6). This is unlikely to be due to poor penetration of drugs since bronchoalveolar lavage sampling of patients on treatment demonstrate sterilizing concentrations of the standard TB drugs in PLF (*44–47*). Alternatively, this aerosolized *Mtb* population may exhibit phenotypic drug-tolerance with clearance from this compartment primarily driven by innate immune processes. The ability of the aerosol phenotype of *Mtb* to persist in bioaerosols during and after TB therapy poses a previously unrecognized challenge to TB eradication within an individual. Furthermore, the isolation of viable *Mtb* from individuals not reaching a clinical threshold for TB diagnosis and treatment potentially expands the cohort of sub-clinical TB in the community. However, whether these organisms are transmissible to other hosts and have the capacity to revert to disease-causing phenotypes remains to be determined.

It is also notable that symptom resolution is independent of treatment (Fig. 5B). The proportion of individuals both with detectable *Mtb* bacilli in sampled aerosol and presence of TB symptoms decline over a similar timescale. This lends support to an immunological PLF clearance hypothesis in which *Mtb* activated macrophages release tumor necrosis factor alpha (TNFα) and other cytokines which play a role in control of *Mtb* infection and at the same time generate systemic symptoms. These findings in the untreated participants (Group C) may indicate transient and spontaneously resolving *Mtb* infection, analogous to the oscillating sub-group originally proposed decades ago (*48*) and reiterated recently (*22*). It remains uncertain, though, if this is due to a new TB exposure or to a perturbation of an existing, stable host-pathogen relationship. Sputum-GeneXpert-negative patients did present with clinical symptoms, but these resolved in parallel with decreasing *Mtb* numbers. DMN-trehalose uptake by bioaerosol-derived *Mtb* indicates that the captured bacilli are metabolically active but does not necessarily provide evidence that this specific phenotype has a role in TB transmission. However, unidentified *Mtb* transmitters, even at low levels, could account for a significant attributable proportion of community exposure exacerbated by long infectious periods and lack of debilitating disease, as highlighted by a recent modelling study (*49*).

Limitations to this study include the pragmatic design (*24*), such that the patient groups were determined by the standard procedures and investigations of South African TB Control Program (*14*). Therefore, the findings reflect a real-world healthcare scenario in South Africa and may lack the precision of a study with more thorough patient investigations – which might include both radiographical and immunological assays. In accordance with the pre-planned pragmatic group allocation (*24*), no additional radiological investigations were carried out as part of this study and the exclusion of TB as likely diagnosis was made by TB clinicians without recourse to routine chest radiography. It is noteworthy, however, that only one individual initially allocated to sputum-GeneXpert-negative untreated group had ongoing symptoms which prompted a subsequent sputum GeneXpert-negative TB diagnosis and assignment to receive TB therapy. Additionally, the limited follow-up of six months gave only a brief snap-shot of the important longer-term relationship between the host and pathogen. It will be important to establish the role that continued *Mtb* infection may have for ongoing transmission, acquisition of new (super)infection and progression to disease. This study was performed in one of the highest TB-burdened populations in the world, so the observations may not be generalizable to other lesser-burdened populations. It is also worth noting that the study was undertaken between June 2020 and June 2022, a period falling within the global Covid-19 pandemic with nationally mandated social interaction constraints and negative impacts on the health infrastructure including TB services.

Our knowledge of the TB host-pathogen relationship has been largely informed over the last 150 years by sputum-based studies. Our understanding of any disease is dependent on the assays and tools which are available to study it. New technologies such as FDG-PET/CT imaging have changed our perspective of within-host TB pathology and developments in non-invasive bioaerosol sampling, such as modified facemasks (*12*) and our RASC sampling system, are revealing new insights in the natural history of TB infection and disease. Applying these technologies longitudinally and to a wide spectrum of at-risk individuals can help to elucidate the complex interactions between pathogen and host throughout the human lifespan. This gives a new window onto individual infectiousness with the potential to inform public health interventions, as well as provide a novel assay with potential utility for assessing targeted chemotherapy and as a metric in vaccine studies.

## MATERIALS AND METHODS

### Study design and population

We recruited consecutive presumptive pulmonary TB patients over the age of 13 who self-presented to two community clinics serving two high-density, peri-urban residential areas south-west of Cape Town, South Africa. A study protocol was published in advance (*24*) with the primary aim to compare proportions of *Mtb*-containing aerosols between sputum-GeneXpert-positive (Group A) and sputum-GeneXpert-negative (Group B) TB patients. Assuming 20% bioaerosol positivity in Group B, and 100% positivity in Group A, a sample size of 250 was calculated to give 90% power to detect a difference between the groups (*24*). As an interim analysis showed no difference in the proportions of positive aerosols between Groups A and B, the study was discontinued after recruitment of 102 patients. Ascertaining the prevalence proportion in Group C was an exploratory aim and the isolation of a high bioaerosol-positive proportion in this group was unexpected. The detection of viable *Mtb* aerosols in the non-TB group, coupled with the concern that they might have unrecognized sputum-GeneXpert-negative TB disease, necessitated longitudinal monitoring and sampling equivalent to the treated TB patients. Sampling intervals of baseline, two weeks, two months, and six months were therefore applied to all three groups, A-C.

### Sampling protocol and data collection

A direct sampling protocol was developed as previously described (*24*) and implemented with slight modifications (*3*). Briefly, the respiratory aerosol sampling chamber (RASC) is a HEPA-filtered enclosure designed for investigation of respiratory bioaerosol emissions from a single individual. The RASC accommodates a Tyvek-suited participant seated and sampled via a metallic elliptical cone which comfortably accommodates each participant’s head (*10*). A unidirectional airflow is created by a high-flow (300L per minute) bespoke cyclone collector connected at the cone apex which extracts bioaerosol into sterile phosphate-buffered saline (PBS). Air enters the collector via a tangential nozzle which generates a liquid cyclone with particle inertia leading to deposition of bioaerosol from the airstream onto the wet wall. The exit airflow from the cone reaches a velocity of 12.5m per second, enabling collection of expiratory aerosols. Seated participants were directed by a study nurse to complete a 15-minute sampling protocol comprising 5-minute sampling for each of 15 Forced Vital Capacity maneuvers (FVCs), tidal breathing, and 15 voluntary coughs . The bioaerosol samples were collected in three separate cones and assessed independently. Ozone sterilization and empty RASC sampling were performed between participants to control for contamination. No *Mtb* bacilli were identified in these empty RASC controls.

At each visit, questionnaires were completed to determine the presence of symptoms including persistent cough (>2 weeks), recent weight loss, night sweats, loss of appetite, fever, hemoptysis, myalgias, and anosmia.

### *Mtb* detection methods

To confirm specificity of *Mtb* detection, both DMN-trehalose and conventional *Mtb* identification methods were employed. Additional assays could not be sequentially applied post DMN-trehalose staining, moreover the paucibacillary nature of the specimen precluded splitting of the sample. Therefore, for subsets of patient visits, additional bioaerosol sampling was performed in parallel to produce a second sample for investigation by Auramine O and/or Ziehl-Neelsen staining and/or DNA analysis using a droplet digital (dd)PCR-based technique.

### Sampling processing

For each participant, bioaerosol material was captured in 5-10mL PBS during each of the three respiratory maneuvers. After centrifugation at 3000 × *g* for 10 min, the pellet was resuspended in 200μL Middlebrook 7H9 medium and stained with the DMN-trehalose probe (*26*). Following overnight incubation in media containing DMN-trehalose, washed samples were added to a nanowell device (*26*). The devices were sealed with adhesive film and centrifuged prior to visualization of the entire array by fluorescence microscopy. Detection and enumeration of fluorescent bacilli was based on DMN-trehalose incorporation and bacterial length and width assessments (*19*) by 2 separate microscopists for each sample, blinded to participants and controls.

### Auramine O and Ziehl-Neelsen Staining

A selection of parallel samples which were not subjected to DMN-trehalose staining were processed for Auramine O and/or Ziehl-Neelsen assays. After overnight incubation in 200μL Middlebrook 7H9 medium, the aerosol pellet was smeared on a slide and microscopy performed using Auramine O fluorescent and/or Ziehl-Neelsen acid-fast staining in accordance with the MGIT procedure manual (*50*). *Mtb* H37Ra smears were prepared as positive controls for the staining techniques.

### *Mtb* RD9 detection and quantification by droplet digital (dd)PCR

Extraction of *Mtb* genomic DNA was accomplished using a modified version of a previously published protocol (51) *Mtb* bioaerosol samples were incubated in 1:1 (v:v) GeneXpert buffer for 15 min, with vigorous shaking every 5 min. The buffer was neutralized by the addition of 1200 ul of dH20 and the sample was centrifuged at 16000 × *g* for 10 min. The supernatant was discarded, the pellet was resuspended in 20μL Tris-EDTA (10 mM Tris, 1 mM EDTA, pH 8·0) and the sample was frozen at -70 °C for 10 min. After thawing at room temperature, the DNA used for ddPCR. The primer/probe combinations and reaction conditions for *Mtb* RD9-specific ddPCR have been described previously (*8*). Serial dilutions of known concentrations of purified *Mtb* H37Rv genomic DNA were included as positive internal controls for ddPCR, and nuclease-free water was included as a negative control. Data generated from the ddPCR reactions were analyzed with the Umbrella pipeline (*52*) using only wells for which a minimum 10,000 droplets were detected.

### *Mtb* whole-genome sequencing

Five samples with the highest number of RD9 copies following 50 days’ incubation of aerosol pellets (n=64) in 200uL Middlebrook 7H9-OADC medium at 37°C were selected for WGS. Prior to library preparation, DNA that had been extracted as described above was purified using AMPure XP magnetic beads as described in (53). Sequencing libraries were prepared using the Nextera XT kit and sequenced on an Illumina NovoSeq platform. WGS was analysed according to previously described methods (54). Briefly, reads were trimmed using Trimmomatic v0.39 (55) with a sliding window of 5:20 and retaining reads with a minimum length of 20 was used to trim reads. Reads were then mapped to the reconstructed ancestor of the MTBC (56) using bwa v0.717 (57) Duplicates were removed using Picard v2.9.1 (58), prior to using Samtools v1.5 (59) and varScan v2.2.4 (60) call variants, with filters to exclude sites with fewer than 10 reads support and minimum base quality scores of 20. The resulting VCF files were used to search for lineage-defining SNPs as described (21) Only lineage-defining SNPs supported by at least 20 reads with an average Phred quality score of at least 30 are reported.

### Fluorescence microscopy

Imaging was performed on a Zeiss Axio Observer 7 equipped with a 100× plan-apochromatic phase 3 oil immersion objective with a numerical aperture of 1.4. Epifluorescent illumination was provided by a 475 nm LED and non-specific fluorescence was removed with a Zeiss 38 HE filter set. Images were acquired using the Zeiss Zen software, and quantitative data extracted using MicrobeJ (*61*).

### Statistical analyses

The bioaerosol positivity proportions of groups A, B, and C were compared using a Fisher’s exact test and *Mtb* numbers with Wilcoxon rank sum tests. Unadjusted odd ratios were used to investigate the impact of baseline demographic and clinical variables on the likelihood of baseline *Mtb* bacillary count positivity. Further logistical regression analyses assessed whether bioaerosol clearance was associated with baseline variables. The three separate timepoints (two weeks, two months, and six months) were investigated for clearance. For each analysis, data were dichotomized into count trajectories that cleared by that visit, defined as “no detectable DMN-trehalose *Mtb* bacilli”, and those that did not clear. Additional analyses used the log-rank statistic for time to *Mtb* clearance and time to resolution of symptoms, defined as the first visit with “no detectable DMN-trehalose *Mtb* bacilli” and with no reported TB screening symptoms respectively. Clearance/resolution proportion for both outcomes were stratified by treatment (groups A and B *vs* group C), previous history of TB disease and HIV status. All statistical analyses were performed using R Core Team (2021) and the R software package, ‘survival’.

### Ethics approval

This study was approved by the Human Research Ethics Committee (HREC/REF: 529/2019) of the University of Cape Town. Written informed consent, including for publication of medical details, was obtained from all participants, and assent was obtained from under 18-year-olds.

### List of Supplementary Materials

**Figure S1:** (**A**) Counts of *Mtb* bacilli at the first visit for all groups, comparing each separate 5-minute respiratory maneuver; (**B**) *Mtb* bioaerosols were released during all three respiratory maneuvers; (**C**) *Mtb* bioaerosols were released during all three respiratory maneuvers.

**Figure S2:** The median time between visits was similar for all three groups: (**A**) The median days at each visit; (**B**) Table indicating the median days until the visit for each group; (**C**) Histogram of visits for all groups.

**Figure S3:** The secular trends in the number of *Mtb* detected in the bioaerosol of each diagnostic group.

**Figure S4.1:** Visit 2 (2-week) clearance of *Mtb* from the bioaerosol for each diagnostic and treatment group stratified by (**A**) diagnostic and (**B**) treatment groups. (**C**) The results of a logistic regression for odds of clearing the bioaerosol at this timepoint

**Figure S4.2:** Visit 3 (2-month) clearance of *Mtb* from the bioaerosol for each diagnostic and treatment group. (**A**) diagnostic and (**B**) treatment groups. (**C**) The results of a logistic regression for odds of clearing the bioaerosol at this timepoint

**Figure S4.3:** Visit 4 (6-month) clearance of *Mtb* from the bioaerosol for each diagnostic and treatment group. (**A**) diagnostic and (**B**) treatment groups. (**C**) The results of a logistic regression for odds of clearing the bioaerosol at this timepoint

**Figure S5:** The length distribution of *Mtb* aerosolized by each respiratory maneuver.

**Figure S6:** Variation in DMN-trehalose staining at baseline.

**Figure S7:** Dimensionality reduction of several morphological and staining characteristics comparing the average bacterial phenotypes from patients on and not on treatment.

## Data Availability

All data produced in the present study are available upon reasonable request to the authors

## Acknowledgments

**Funding:** RW discloses support for this work from the South African Medical Research Council [MRC-RFA-UFSP-01-2013/CCAMP] and the National Institute of Allergy and Infectious Diseases of the US National Institutes of Health under award number R01AI147347 and through the Myco3V Tuberculosis Research Unit (U19AI162584). DFW acknowledges the support of the Strategic Health Innovations Partnerships (SHIP) Unit of the South African Medical Research Council with funds from National Treasury under its Economic Competitiveness and Support Package, and is grateful for funding from the Research Council of Norway (R&D Project 309592). **Author contributions:** Conceptualization: RW, DFW, BP, RD; Methodology: RW, DFW, RD, SG, AK, RS; Investigation: ZH, VJ, BL, AM, RS, AV; Visualization: BP, RD, RW; Funding acquisition: RW, DFW; Project administration: RS, AV, RW; Supervision: RW, DFW, SH, FC; Writing – original draft: BP, RD, DFW, RW, AK; Writing – review & editing: BP, RD, DFW, RW, AK, SG, FC, SH. **Competing interests:** The authors declare that they have no competing interests. **Data and materials availability:** All data are available in the main text or the supplementary materials.

## Notes

### Competing Interest Statement

The authors have declared no competing interest.

### Author Declarations

This study was approved by the Human Research Ethics Committee (HREC/REF: 529/2019) of the University of Cape Town.

### Summary of Updates

New analysis exploring the variables associated with clearance of aerosolized Mtb and loss of symptoms over time. New data concerning the microscopic characterization of Mtb bacilli from bioaerosol samples including change in DMN-trehalose phenotype and Mtb cell length over time. Revised discussion. Revised figures and supplementary material.

