## Supplemental Figures and Tables for "Aerosolization of viable *Mycobacterium tuberculosis* bacilli by tuberculosis clinic attendees independent of sputum-GeneXpert status": ABC_Supplementary Material.pdf

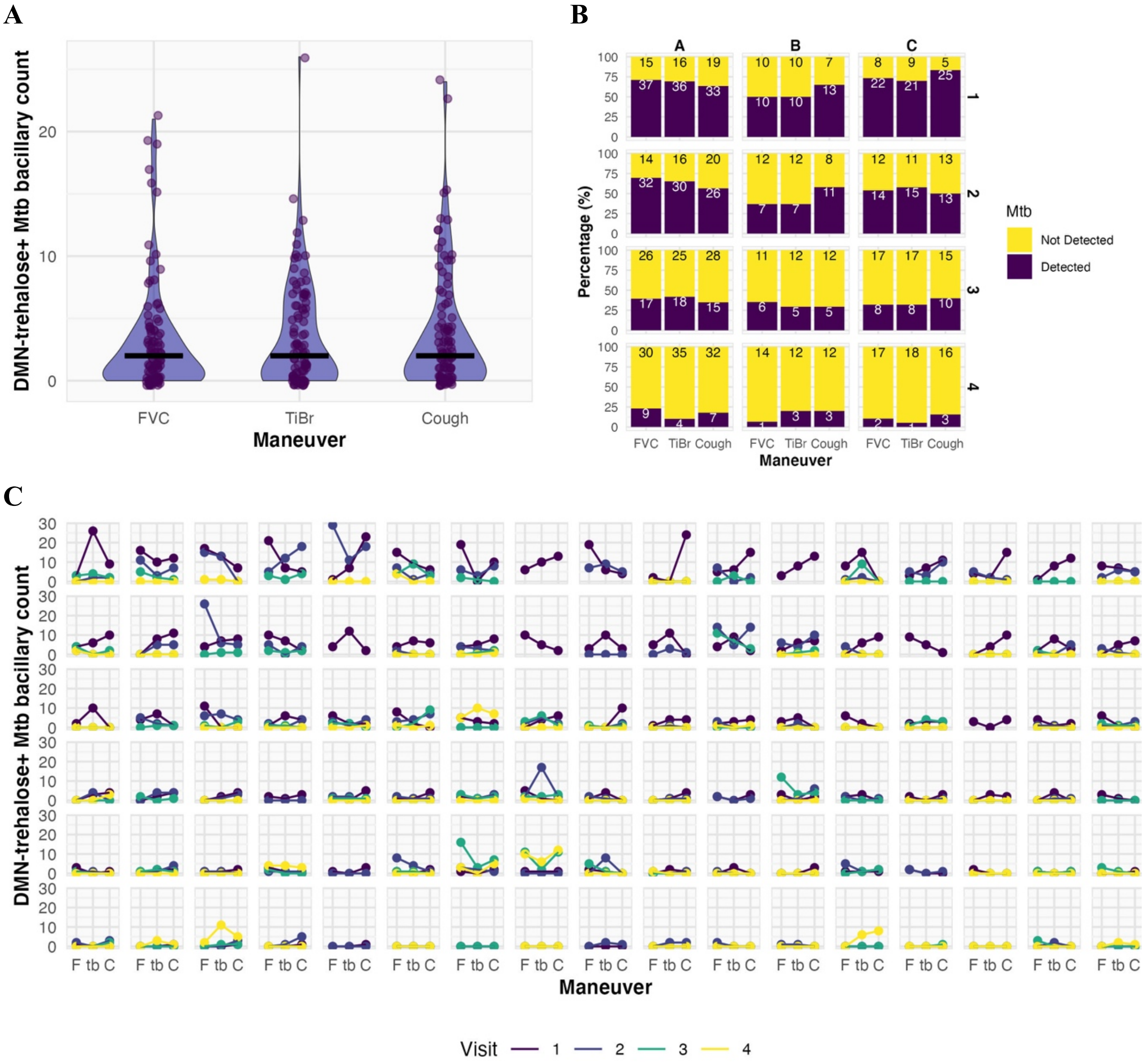


**Fig. S1.** *M. tuberculosis* (*Mtb*) bioaerosols were detected in all three diagnostic groups, from all respiratory maneuvers, and at all visits. (**A**) Counts of *Mtb* bacilli at the first visit for all groups, comparing each separate 5-minute respiratory maneuver. The maneuvers are compared with thick bars representing median values. Wilcoxon Signed Rank test found no significant differences between the bacillary output for each maneuver after Bonferroni correction for multiple comparisons. (**B**) The percentage of bioaerosol samples that were positive for *Mtb*, grouped by visit (vertical) and diagnostic group (horizontal). Fisher's exact tests performed comparing each group at each timepoint found no significant differences after correction for multiple comparison. FVC, forced vital capacity; TiBr, tidal breathing. (**C**) The number of putative *Mtb* detected per respiratory maneuver stratified by visit. After grouping for visit and diagnostic group, the average number of *Mtb* per maneuver was compared using a Wilcoxon Signed Rank test, followed by a Bonferroni correction for multiple comparisons. No significant differences were detected. F, forced vital capacity; tb, tidal breathing; C, cough.

**A**

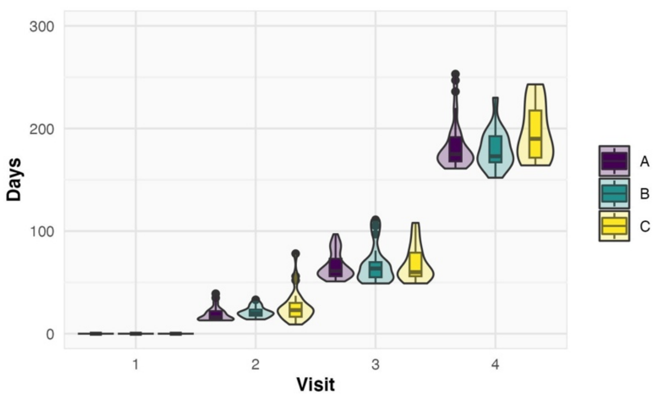


**B**

| Group | Visit | Median | Lower Quartile | Upper Quartile |
|---|---|---|---|---|
| A | 2 | 16 | 14 | 22 |
| B | 2 | 20 | 18 | 24 |
| C | 2 | 23 | 17 | 30 |
| A | 3 | 61 | 56 | 73 |
| B | 3 | 63 | 55 | 69 |
| C | 3 | 60 | 56 | 79 |
| A | 4 | 175 | 168 | 192 |
| B | 4 | 172 | 167 | 188 |
| C | 4 | 190 | 172 | 218 |

**C**

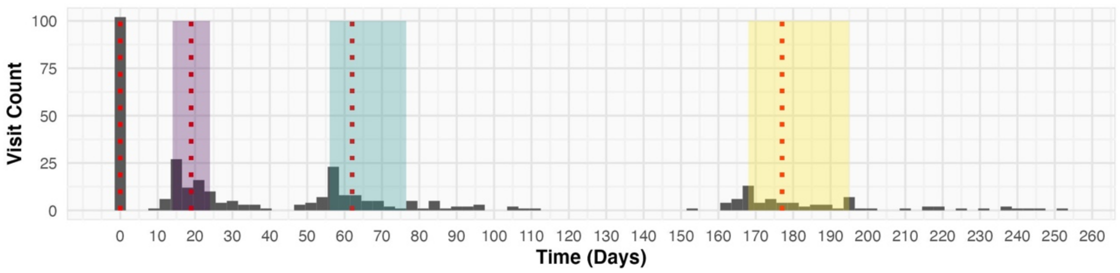


**Fig. S2. The median time between visits was similar for all three groups. (A)** The median days at each visit was compared between the three groups using a Wilcoxon Rank Sum test, followed by a Bonferroni correction for multiple comparisons. No significant differences were detected. **(B)** Table indicating the median days until the visit for each group. **(C)** Histogram of visits for all groups. Interquartile time window highlighted for each visit, dotted red line represents the median timepoint.

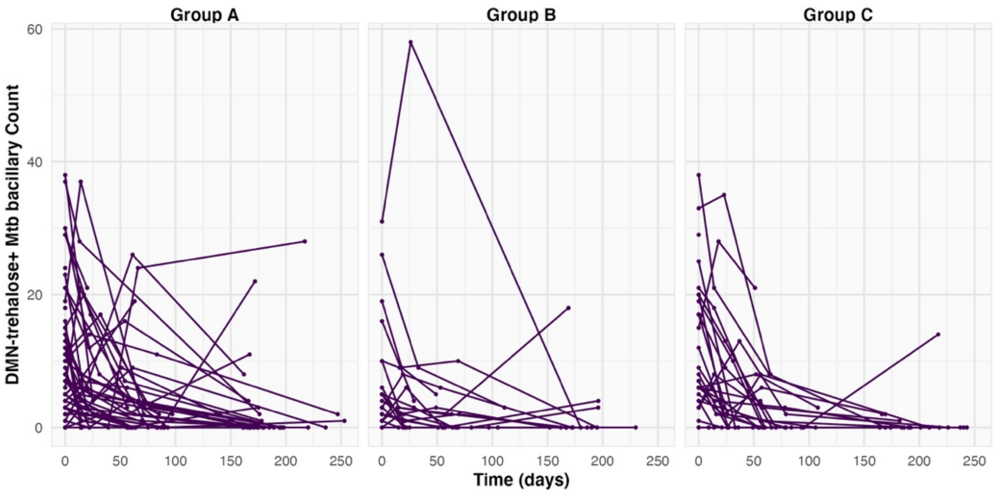


**Fig. S3. Secular trends in the number of *Mtb* detected in bioaerosol samples of each diagnostic group.** Lines connect the samples from the same participant at each successive visit.

**A** **B**

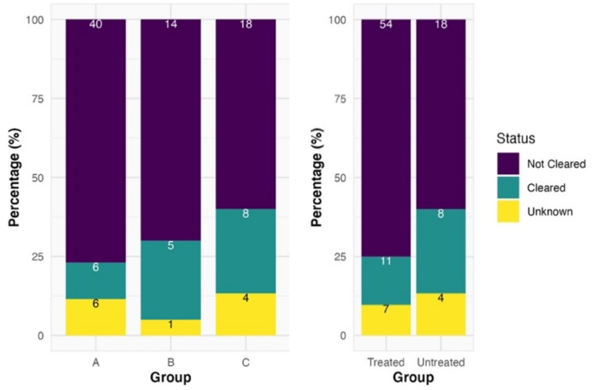


**C**

| | Odds Ratio | Low CI (2.5) | High CI (97.5) | p-value |
|---|---|---|---|---|
| **Age (≥30)** | 0.27 | 0.05 | 1.31 | 0.113 |
| **Sex (Male)** | 4.09 | 1.07 | 19.97 | 0.054 |
| **HIV Status (Positive)** | 1.44 | 0.42 | 5.14 | 0.564 |
| **Body Mass Index (≥20)** | 0.85 | 0.26 | 2.74 | 0.779 |
| **Previous History of TB** | 2.62 | 0.77 | 9.8 | 0.131 |
| **GeneXpert (Positive)** | 0.43 | 0.08 | 2.19 | 0.309 |
| **Treated** | 0.57 | 0.12 | 2.42 | 0.452 |

**Fig. S4.1: Visit 2 (3-week) clearance of *Mtb* from the bioaerosol for each diagnostic and treatment group.** The percentage of participants in which the bioaerosol was “not cleared”, “cleared” or “unknown” (due to a missed visit) is shown stratified by **(A)** diagnostic and **(B)** treatment groups. **(C)** The results of a logistic regression of baseline characteristics with the odds of clearing the bioaerosol at this timepoint. For this analysis, individuals defined as “unknown” were removed.

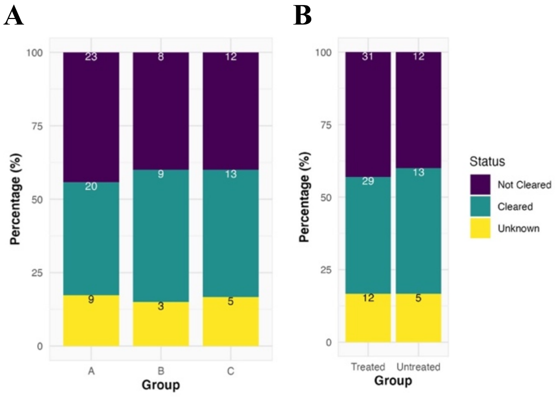


**C**

| | Odds Ratio | Low CI (2.5) | High CI (97.5) | p-value |
|---|---|---|---|---|
| **Age (≥30)** | 2.25 | 0.69 | 7.8 | 0.185 |
| **Sex (Male)** | 0.61 | 0.21 | 1.6 | 0.332 |
| **HIV Status (Positive)** | 0.56 | 0.19 | 1.6 | 0.275 |
| **Body Mass Index (≥20)** | 0.52 | 0.18 | 1.4 | 0.189 |
| **Previous History of TB** | 0.88 | 0.30 | 2.6 | 0.822 |
| **GeneXpert (Positive)** | 0.66 | 0.16 | 2.6 | 0.546 |
| **Treated** | 1.44 | 0.36 | 5.9 | 0.606 |

**Fig. S4.2: Visit 3 (2-month) clearance of *Mtb* from the bioaerosol for each diagnostic and treatment group.** The percentage of participants in which the bioaerosol was “not cleared”, “cleared” or “unknown” (due to a missed visit) is shown stratified by **(A)** diagnostic and **(B)** treatment groups. **(C)** The results of a logistic regression of baseline characteristics with the odds of clearing the bioaerosol at this timepoint. For this analysis, individuals defined as “unknown” were removed.

**A**

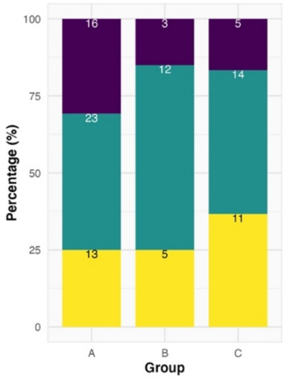


**B**

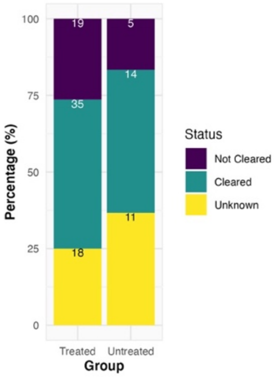


**C**

| | Odds Ratio | Low CI (2.5) | High CI (97.5) | p-value |
|---|---|---|---|---|
| **Age ≥30** | 0.78 | 0.21 | 2.70 | 0.694 |
| **Sex (Male)** | 1.23 | 0.41 | 3.72 | 0.710 |
| **HIV Status (Positive)** | 1.03 | 0.31 | 3.42 | 0.957 |
| **Body Mass Index ≥20** | 1.14 | 0.37 | 3.60 | 0.819 |
| **Previous History of TB** | 0.96 | 0.27 | 3.36 | 0.949 |
| **GeneXpert (Positive)** | 0.31 | 0.06 | 1.43 | 0.153 |
| **Treated** | 1.94 | 0.36 | 11.95 | 0.448 |

**Fig. S4.3. Visit 4 (6-month) clearance of *Mtb* from the bioaerosol for each diagnostic and treatment group.** The percentage of participants in which the bioaerosol was "not cleared", "cleared" or "unknown" (due to a missed visit) is shown stratified by **(A)** diagnostic and **(B)** treatment groups. **(C)** The results of a logistic regression of baseline characteristics with the odds of clearing the bioaerosol at this timepoint. For this analysis, individuals defined as "unknown" were removed.

**A**

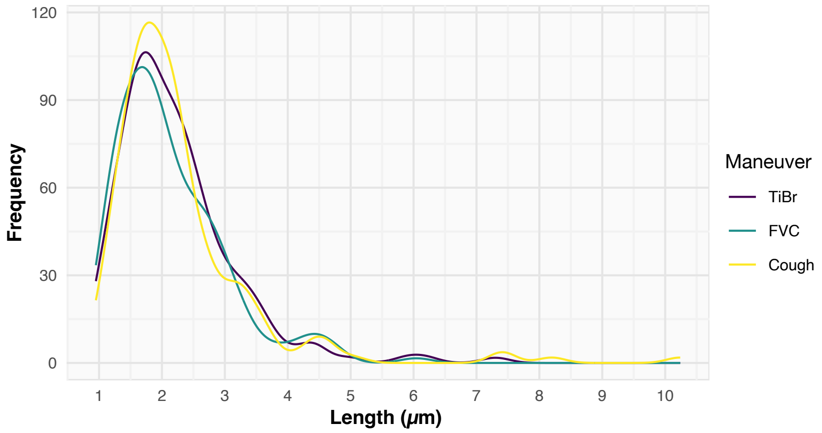


**B**

| | Fold change | 95% CI |
|---|---|---|
| **FVC** | 1.030 | 0.951; 1.117 |
| **Cough** | 0.997 | 0.926; 1.073 |
| **Group B** | 0.970 | 0.726; 1.295 |
| **Group C** | 0.968 | 0.852; 1.100 |

**Fig. S5. The length distribution of *Mtb* aerosolized by each respiratory maneuver. (A)** The length distribution of aerosolized *Mtb*, stratified by respiratory maneuver. **(B)** Results of a linear mixed effects model highlighting that no differences were detected, even when stratified by diagnostic group.

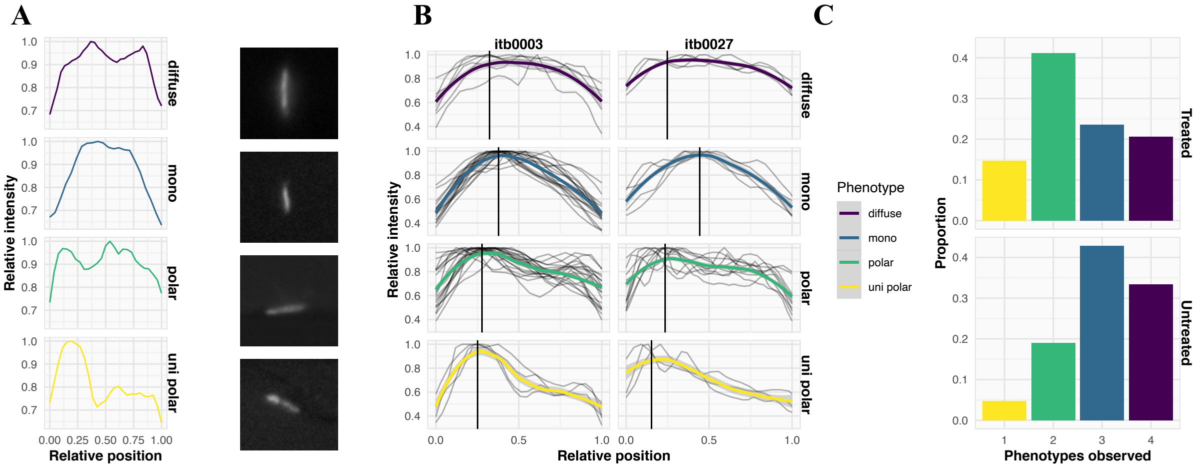


**Fig. S6. Variation in DMN-trehalose staining at baseline. (A)** Representative plots and images indicating the differing distributions of DMN-trehalose along the medial axis of the bacilli. **(B)** Two participants aerosolizing several bacilli with across all the observed phenotypes. **(C)** The proportion of patients at baseline aerosolization *Mtb* with 1-4 of the phenotypes from **(A).**

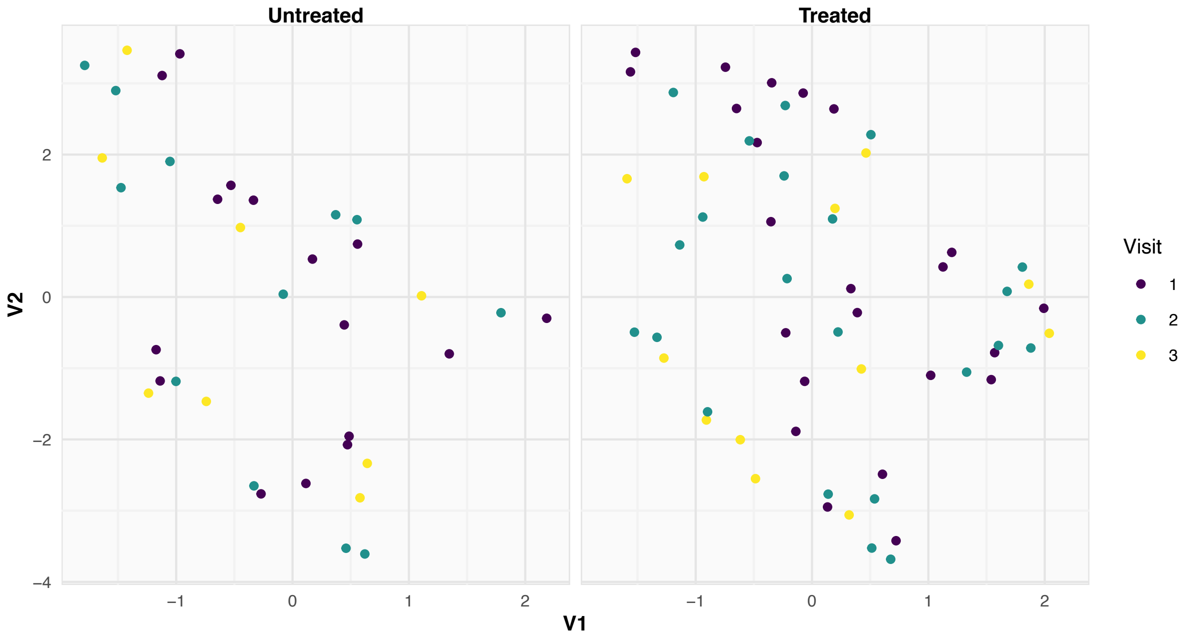


**Fig. S7. Dimensionality reduction of morphological and staining characteristics.** Comparison of average bacterial phenotypes from participants not undergoing standard anti-TB chemotherapy (Untreated) and those on TB treatment (Treated).
